# RSI model: COVID-19 in Germany Alternating quarantine episodes and normal episodes

**DOI:** 10.1101/2020.05.01.20075754

**Authors:** Carolin Knoch, Timo Hartmann, Alan Wildblood

**Affiliations:** TU Berlin

## Abstract

This paper was developed in the Civil Systems Engineering Department at the Technical University of Berlin. The background is the spread of COVID-19 pandemic in Germany. Our current situation is a threat from the COVID-19 virus without vaccine and medication. Germany has increased its intensive capacities, intensified research in its analyzes and research projects and is currently testing the influence of various measures on the rate of expansion.

The goal now is to find a strategy that slows the spread so that medical capacities are not overloaded. The approach is to alternate between quarantine and normal episodes and the result is an oscillation in the number of cases between two limits.

The mathematical model is an SRI model that can be used to calculate the development of the case numbers. All uncertain parameters are varied at significant intervals. Influence parameters and control parameters were defined. By adjusting the length of the episode to match the speed of spread of the virus, the gap can be bridged until a vaccine has been developed or sufficient immunity is available in the population. By observing the capacity limits in connection with the number of cases, the need for a new quarantine episode and the possibility of initiating a normal episode can be predetermined. This does not exceed the capacity limit.

In quarantine, contagion within households is limited by the size of the household; only system-critical professions continue to work in public. All others must remain in the homeschooling / home office. In the normal episode, everyone can work, go to school, have social contacts. A solution could be found in all parameter combinations. The ratio of the normal episode duration / quarantine episode lies between 0.3 and 0.95 in the examination area.

## Introduction

Pandemic containment is essential to prevent overloading of the medical care system. Limitation of the pandemic is mainly facilitated by reducing infections. An effective strategy is social distancing. However, this strategy draws more and more criticism, in Germany as elsewhere, because of its economic and psychological consequences for society. How can the number of infected person be held below the medical care capacity while restrictions affecting the general public are minimized?

Various approaches exist to retarding the infection rate during the current coronavirus (COVID-19) outbreak until vaccine has been developed, without overloading medical capacity. Options being discussed include large-scale testing, contact tracing and social distancing.

A study published by the Imperial College contains a model calculation of the effects of alternating school closings and openings in the United States and the United Kingdom. However, no satisfactory solution was developed. Variables which describe Germany’s medical care resources and the age and occupational structure of its population contrast sharply with American and British conditions.

This study attempts to apply the Imperial College approach of alternating quarantine and normal episodes for the entire population.

Mathematically, an oscillation in hospitalization can be imposed. The difficulty lies in determining the non-controllable parameters related to the virus. Varying the ambiguously quantified input variables is investigated in an attempt to devise a strategy for all combinations that does not overload the clinical system.

## Research Method

To investigate the problem, an SIR model was developed in which quarantine and normal episodes alternate (S = sick, I = infected, R = immune/dead). The modeling also includes infectiousness by forming subgroups in the infected ones.

There can be formed three groups for those who are currently ill. The three groups are based on the mean course of the disease defined, which is shown in red in the right image. Accordingly, there are asymptomatic, symptomatic and not contagious phases exist and thats also what the subgroups define.

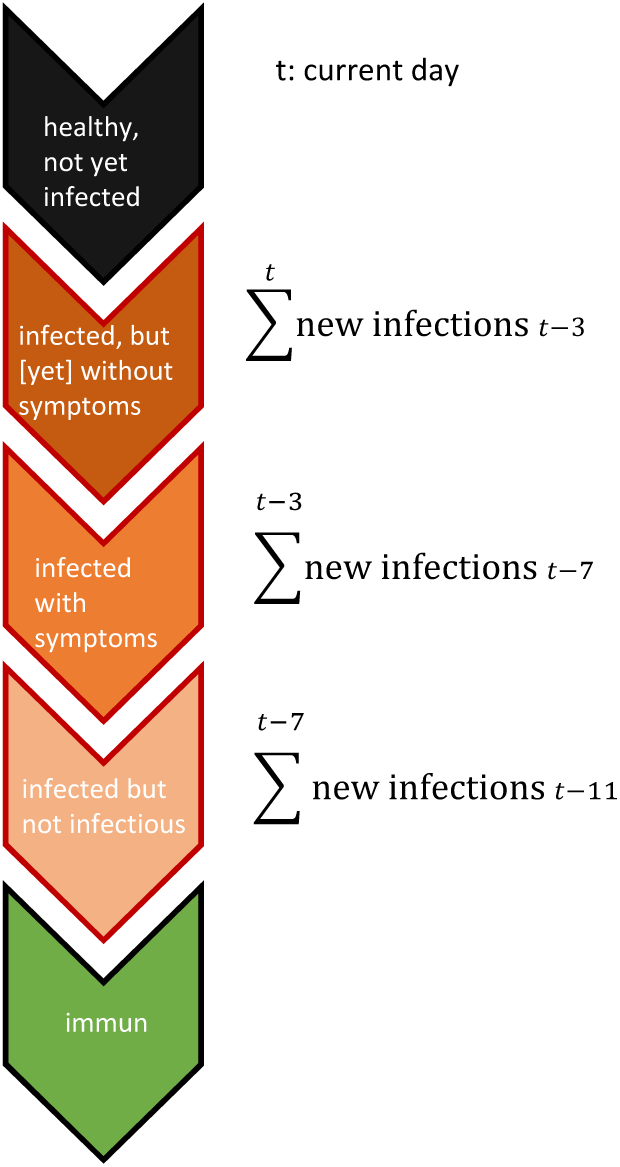

The infection rate in the normal episodes is derived from the number of cases identified in Germany. In the quarantine episodes it is assumed that only people in essential occupations work in public. Contacts are limited to people in their own homes. Thus, the transmission figure is limited in the quarantine episode to the number of persons living in one household.

The uncertainty in these factors is taken into account by varying each independent variable at significant intervals.

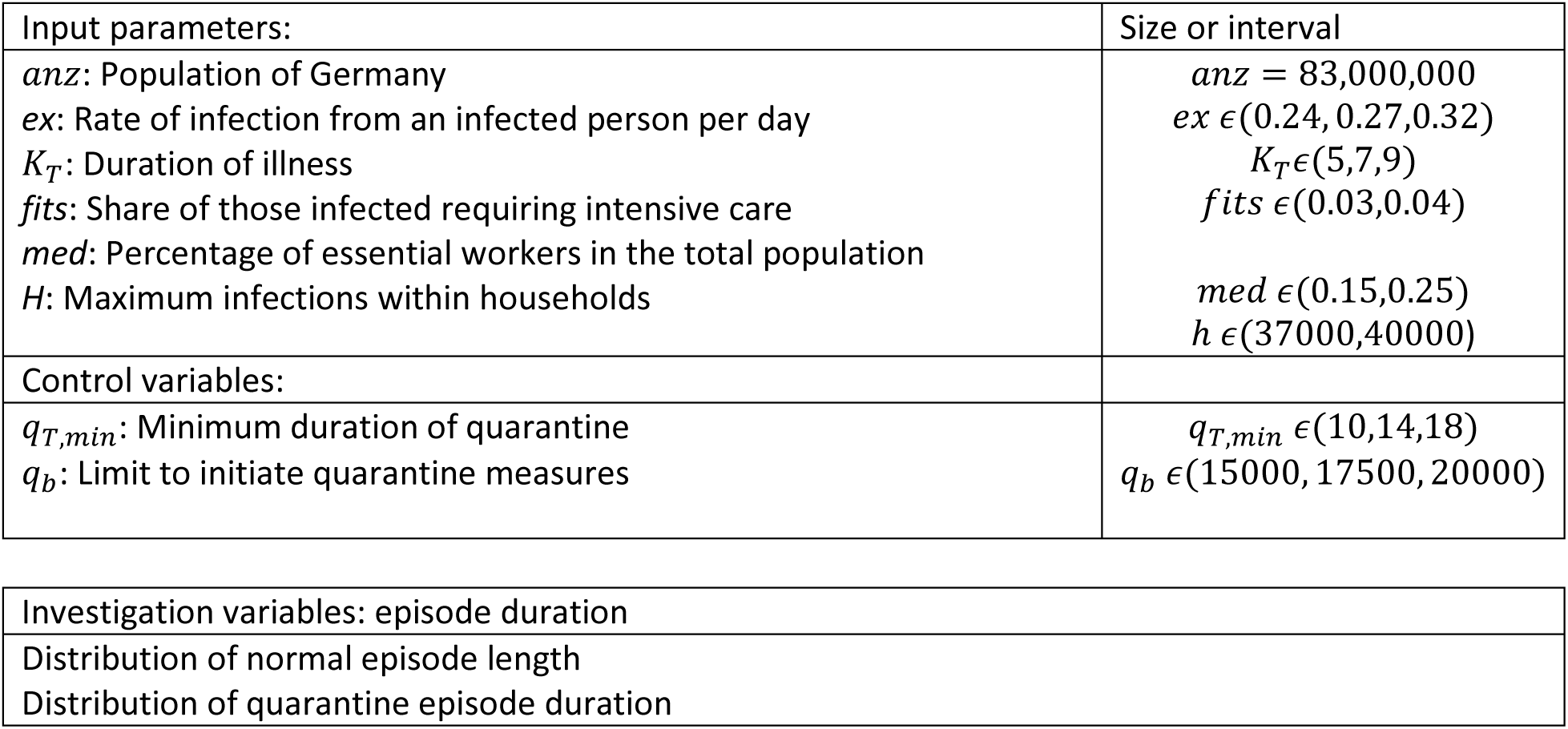

| Input parameters: | Size or interval |
| --- | --- |
| $anz$: Population of Germany<br>$ex$: Rate of infection from an infected person per day<br>$K_T$: Duration of illness<br>$fits$: Share of those infected requiring intensive care<br>$med$: Percentage of essential workers in the total population<br>$H$: Maximum infections within households | $anz = 83,000,000$<br>$ex \in (0.24, 0.27, 0.32)$<br>$K_T \in (5, 7, 9)$<br>$fits \in (0.03, 0.04)$<br>$med \in (0.15, 0.25)$<br>$h \in (37000, 40000)$ |
| Control variables: |  |
| $q_{T,min}$: Minimum duration of quarantine<br>$q_b$: Limit to initiate quarantine measures | $q_{T,min} \in (10, 14, 18)$<br>$q_b \in (15000, 17500, 20000)$ |

|  |
| --- |
| Investigation variables: episode duration |
| Distribution of normal episode length |
| Distribution of quarantine episode duration |

All variations of the control variables are used to generate a solution for each input parameter combination. The quarantine period is automatically extended if the capacity limit would be exceeded again in less than 5 days.

This study provides an overview of possible development scenarios in different situations and the impact of control variables on the rate of spread.

## Data Analysis

### Input parameters

Below, calibration of the input parameters is made. In this model duration of disease is of primary importance for the number of new infections by one infected person. For duration of infectivity 0 to 8 days is chosen in accord with the RKI website.^1^

The infection rate *ex* is the average number of new infections per day per infected person. Multiplied by the duration of participation of infected people in public life, it yields the basic reproduction number *R*_0_. This result depends on how many contacts the infected person has and how long he or she participates in public life without symptoms. The latter depends on highly variable disease progression and the incubation period. In contrast to the daily rate of infection, the basic reproduction number cannot be ascertained directly from the number of cases. Therefore, the duration of infectivity *k_T_* and the rate of infection *ex* can be varied separately in the model.

The infection rate in times without boundaries of society can be derived from the number of cases:

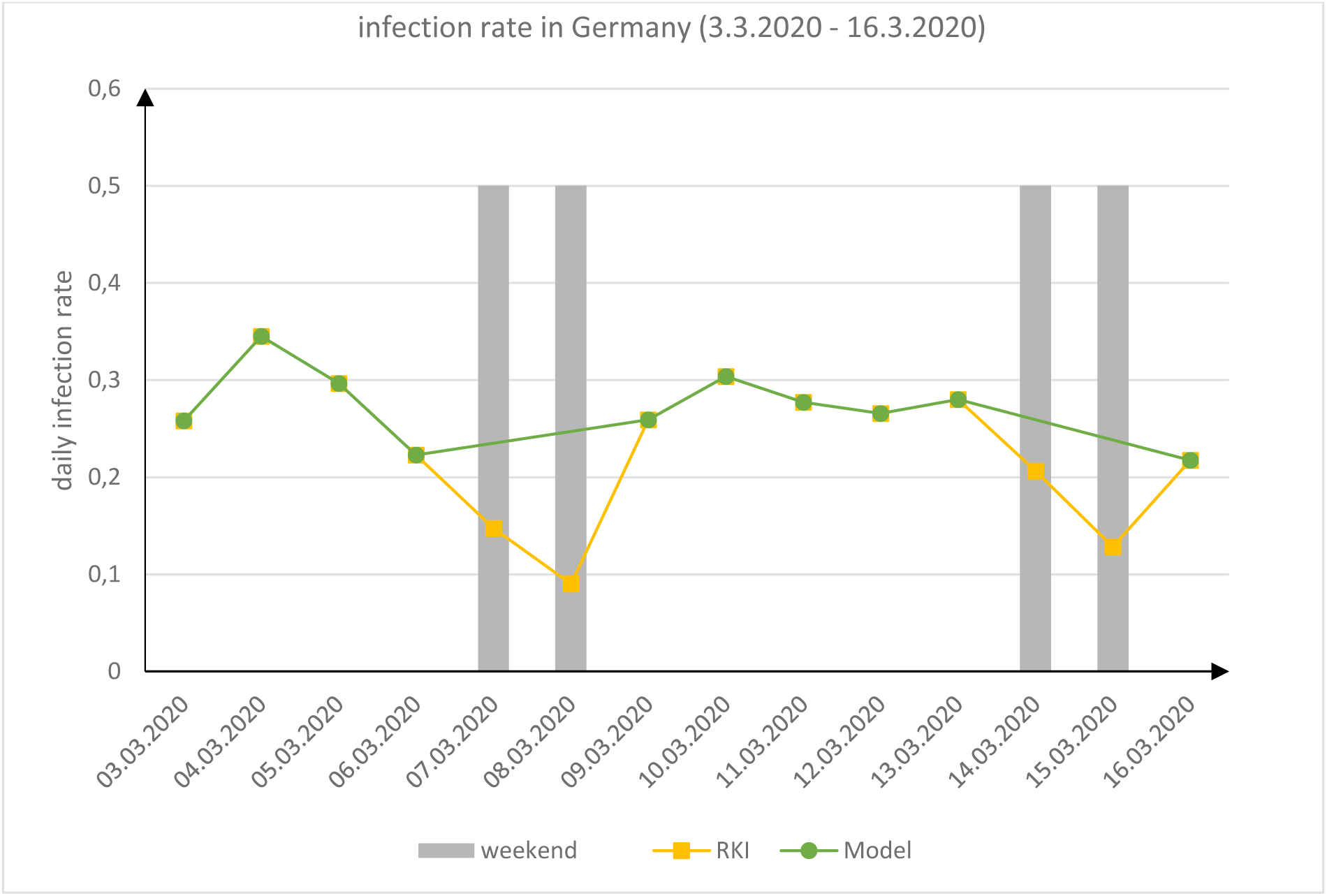

The daily increase in the number of cases as specified by the RKI varies from 0.1 to 0.36. The weekends are marked in gray. Evaluation did not consider these rates because they are clearly lower than the rest. The average value without these rates is thus *ex=0.24*. This greatly overestimates the information *R*_0_ ϵ (2, 3.3) provided by the RKI^1^, but corresponds to the number of cases in early stages of the pandemic.

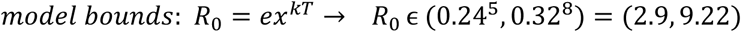

> *Update: 04.05.2020: the number of tests per day and the number positive results were published in the RKI bulletin On 02.04..^2^Therefore, an increase in the number of tests from day to day produces an overestimation of daily increase. This bias has to be adjusted*.

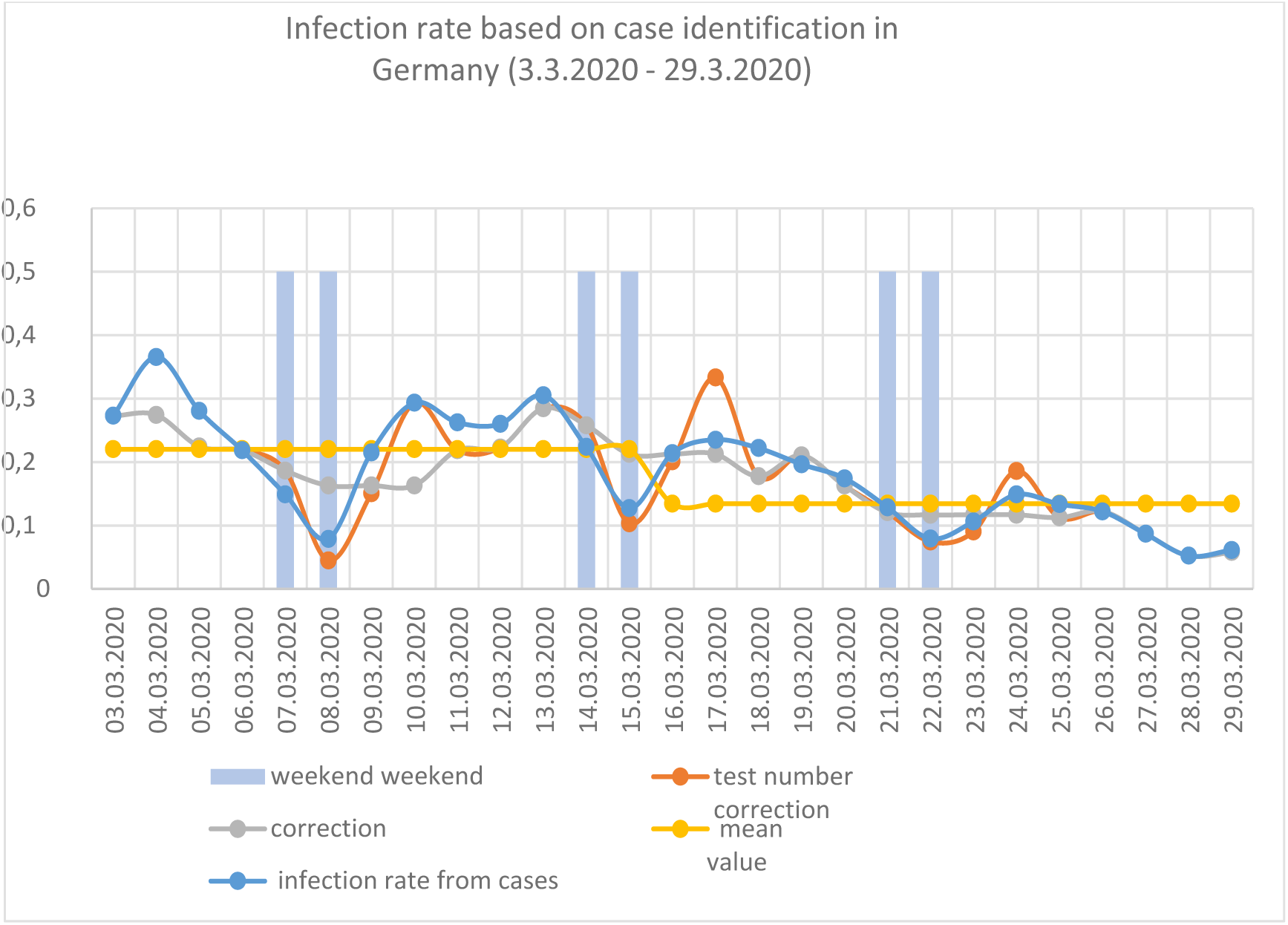

> *The result of the test number correction is factored out as a red line. A further weekend correction yields a balanced growth factor. An average of ex = 0.22 is obtained for the period without measures and a value of ex = 0.13 for the period since restrictions were imposed in Germany*.

An Imperial College study shows the share of different age groups requiring intensive care (IC)^3^. Based on Germany’s age structure^4^, the IC patient proportion *fits* was estimated. The relevant data can be found in Appendix A1.

Even during quarantine episodes people have to work in essential industries in interaction with the public. These vital industries primarily focus on food supply and healthcare. This interaction can cause further infections even during quarantine periods. In Appendix A2 the share *med* of Germans who work in such areas is calculated.

Alternating between episodes has a crucial advantage. The spread is slowed down and there are fewer new cases, sick people become healthy during the quarantine and at the end of each quarantine episode the positive tested infected remain in quarantine over the normal episode. This means that by changing the episodes, phases are created in which all and with them the symptom-free infected of this time are isolated. These Infectivity while being asymptomatic can be partially eliminated by quarantine episode. A representation of a case number development using this model is shown in the figure below. The calculation of the future contains sums over the time of new infections and the number of patients in the past, so that a simpler model is used. It overestimates the increase in the normal episode and the decrease in the quarantine episode. Note the time shift. The new quarantine episode must be initiated in advance before the capacity limit is reached. When calibrating the simple mathematical model, there are only 40000 new infections in the entire quarantine episode.

Calculation model for quarantine episodes:

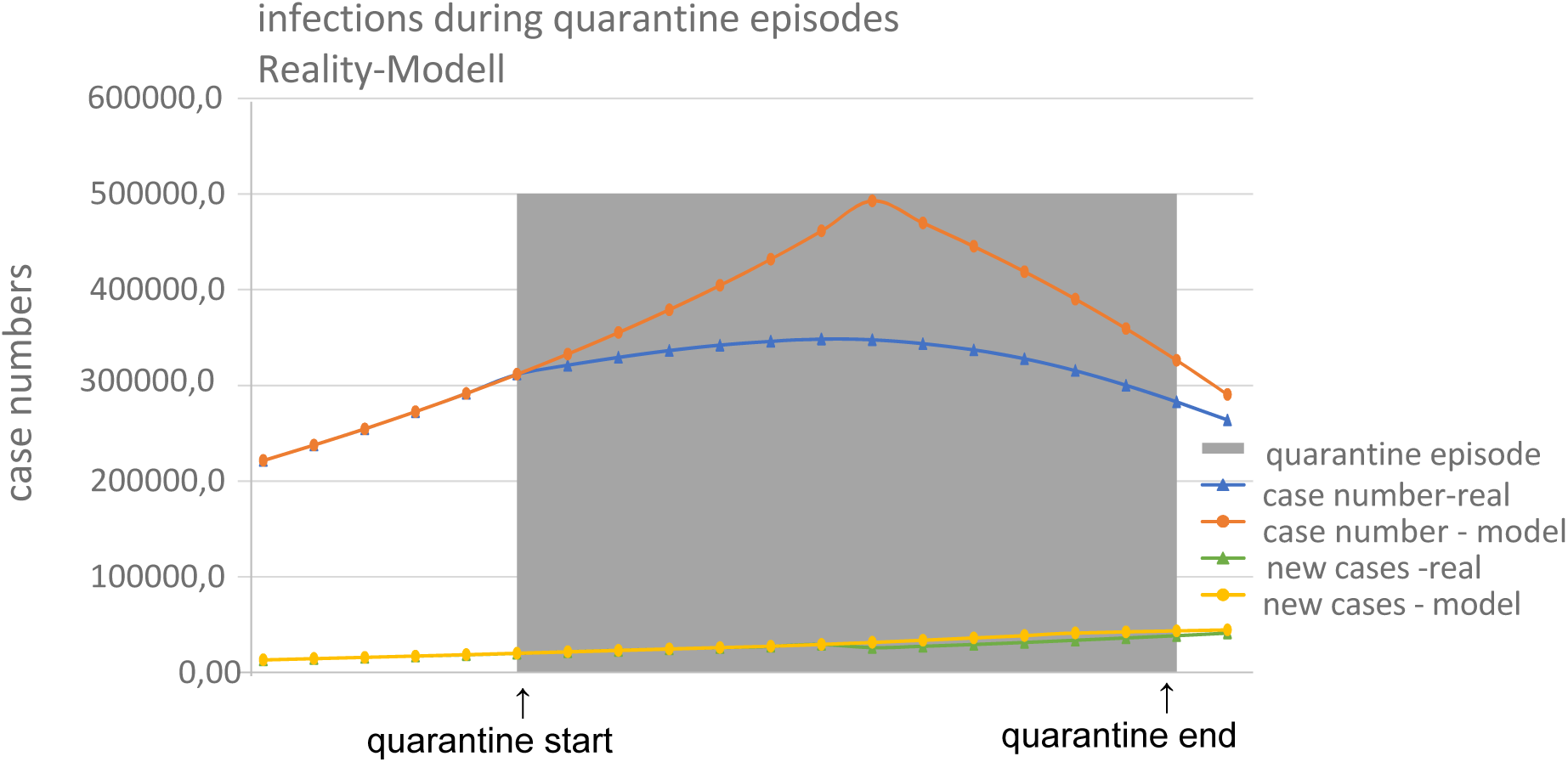

Initiation of a quarantine episode is subject to monitoring IC capacities. In this model, it is linked to a specific anticipated number of patients requiring intensive care. Minimum quarantine duration and the threshold triggering a quarantine episode are varied to determine what influence they have on episode duration.

### Resulting data

From each possible parameter combination, selection of an optimum control variable and scheduling of quarantine and normal episodes are determined as follows. Below, the parameter combination with the shortest normal episodes in relation to the mean quarantine duration is shown. It is parameter combination 642, the combination with the least favorable interval value:

| No. | $ex$ | $k_T$ | $fits$ | $med$ | $h$ |
| --- | --- | --- | --- | --- | --- |
| 642 | 0.32 | 9 | 0.04 | 0.25 | 40000 |

The following control variables were selected from among the nine control variants while monitoring the resulting episode duration ratios:

| $q_{T,min}$ | $q_b$ |
| --- | --- |
| 10 | 20000 |

This results in the following schedule and in A3 more information:

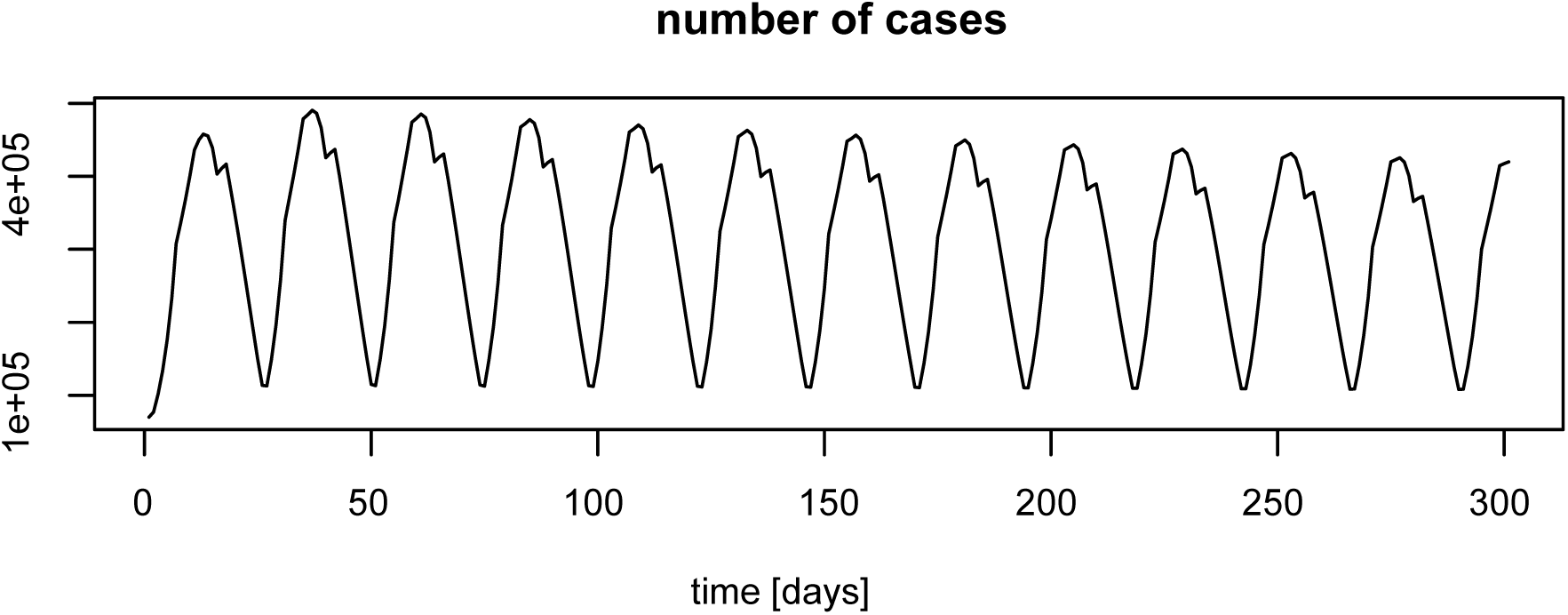

The total infected population ranges from a maximum of 490,700 and a minimum of 70,000. With a 4% share of patients requiring intensive care, these figures vary between 19,625 and 3500. The requirement not to exceed *q_b_*=20000 is met. A total population timeline gives the immune population shown in green, the infected in red and the uninfected in black. In the figure below, the duration of the alternating episodes can be ascertained. On the right a histogram shows the distribution of normal episode (green) and quarantine episode (black) durations.

The quarantine episodes had to be extended from the minimum period *q_T_*_,min_ to 19 days. The normal episodes can be from 5 to 6 days.

The result is a ratio of quarantine duration to normal duration of 0.26.

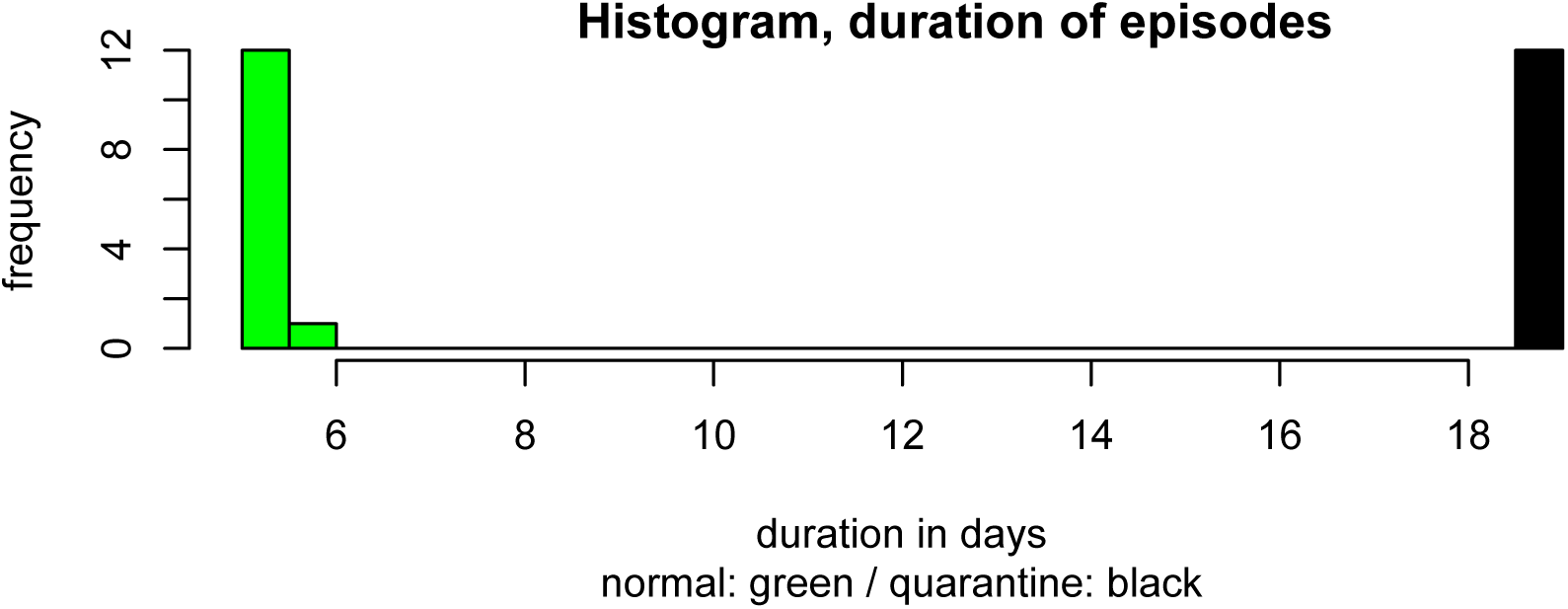

The longest ratio of the average episode durations is obtained with parameter combination 3:

| No. | $ex$ | $k_T$ | $fits$ | $med$ | $h$ |
| --- | --- | --- | --- | --- | --- |
| 3 | 0.24 | 5 | 0.03 | 0.15 | 37000 |

The following control variables were selected for this combination of control parameter

The following control variables were selected for this combination of control parameters :
| $q_{T,min}$ | $q_b$ |
| --- | --- |
| 10 | 20000 |

The results are shown in A4 and in the figures below:

The total number of infected persons ranges between a maximum of 480100 and a minimum of 70,000. With a 4% share of the infected requiring intensive care, they number between 13125 and 3500 persons. The requirement not to exceed *q_b_*=20000 is again met.

The figure below depicts the alternation of quarantine and normal episodes. The quarantine episodes are always 10 days. The normal episodes could be 9–12 days. The duration of normal episodes becomes longer over time because of the growing immunity in the population.

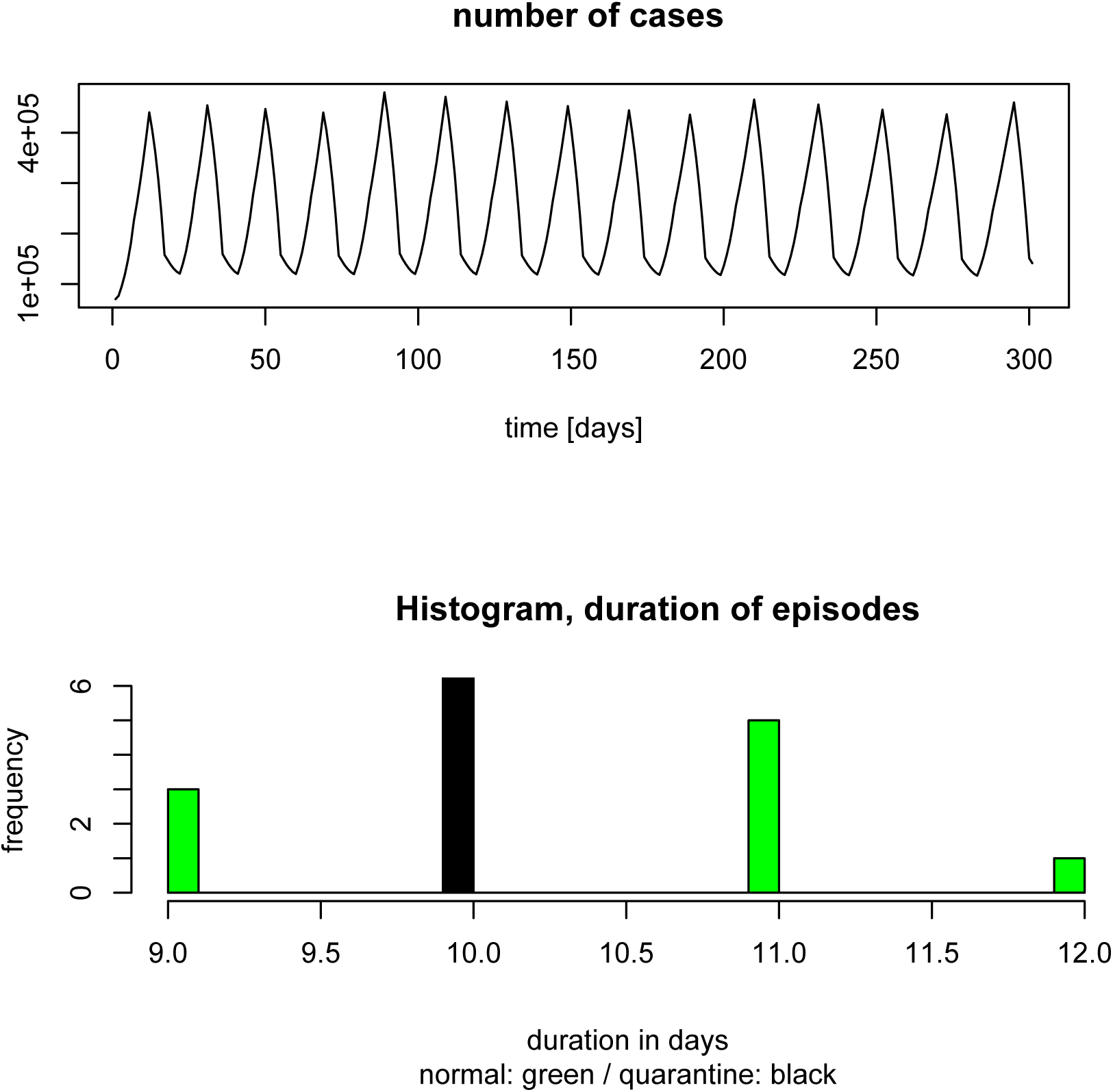

An overall view of the influence of individual parameters (control variables and input parameters) on the average normal episode duration for each solution is provided here by a projection on the appropriate level in the parameter space.

The strong influence of factors ex and *k_T_* is easily recognized. On the contrary, the influence of *fits*, *med* and *h* is negligible. Selection of too short a quarantine period *q_T,min_*leads to a longer normal episode. If the capacity limit *q_b_* is increased, so is the normal episode length.

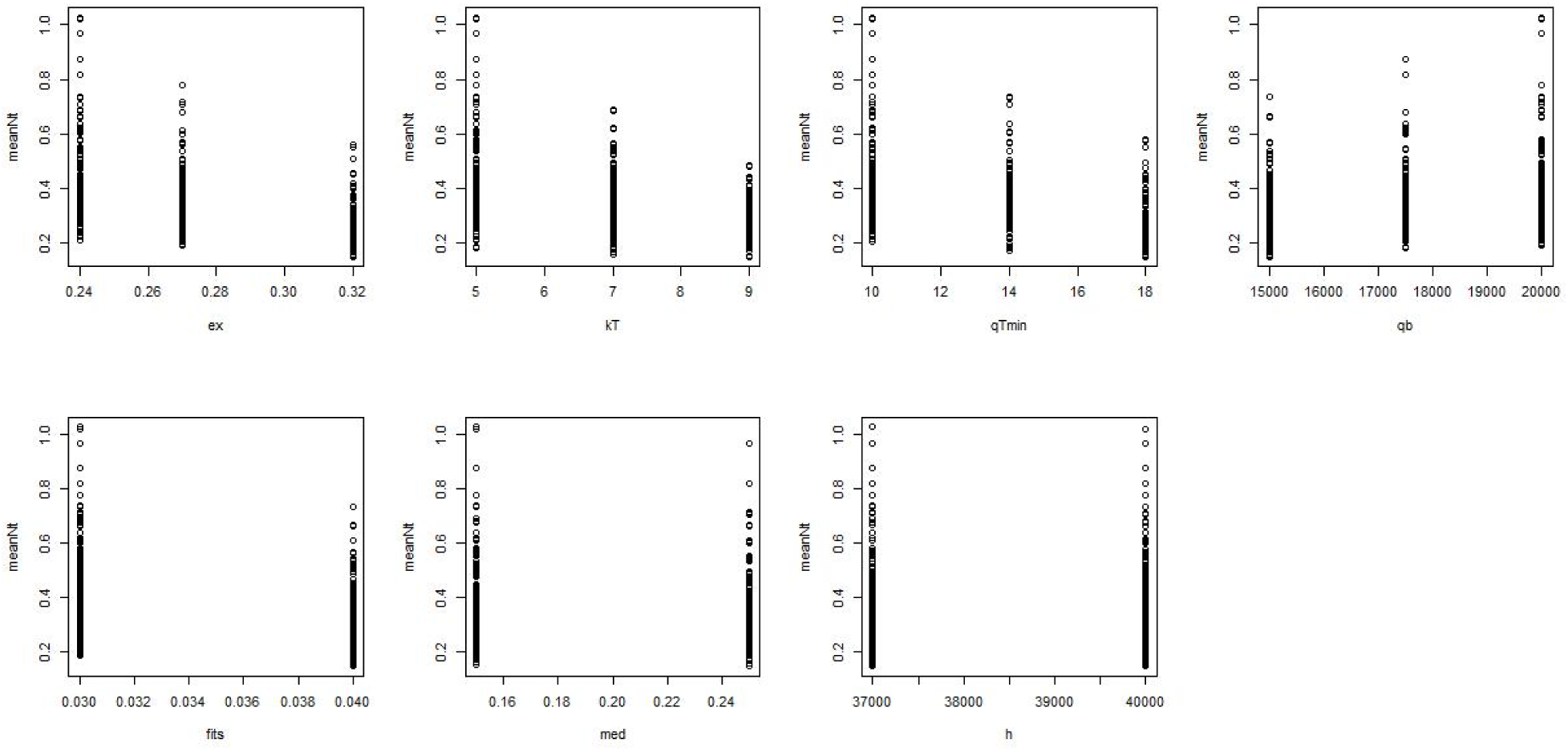

## Discussion and Conclusions

The spread of SARS-CoV-2 requires action. Since flattening below the capacity of the health system is only possible through measures that can be maintained over an extended period, the possibility of alternating shorter episodes with restrictions and normal periods must be investigated.

It is important for compliance with the measures that they be socially accepted. This study is motivated by the assumption that restrictions are more likely to be accepted if they are temporary and interspersed with normal episodes.

> *Update 05.04.2020: Since the interval for the rate of infection in this study is significantly overestimated due to the number of cases in the raw data, the actual rate in realistic scenarios is on the whole better than depicted here*.

No universal procedure can be found for alternating quarantine and normal episodes. This is impossible due to the uncertainty in the model parameters. Nevertheless, a strategy was found for each combination. Thus, this strategy requires constant monitoring of the number of cases and intensive care bed utilization. A new quarantine episode can thus be initiated at any time. Timing the next episode alternation is constantly improved thanks to availability of data from past episodes.

No assessment of this strategy compared to other strategies is made in this study. It is simply one possible approach among many to bridge the gap until a vaccine can be developed.

## Ongoing investigations

Another model being developed. Instead of alternating episodes in the total population, the population is divided into two groups, as shown in the following figure:

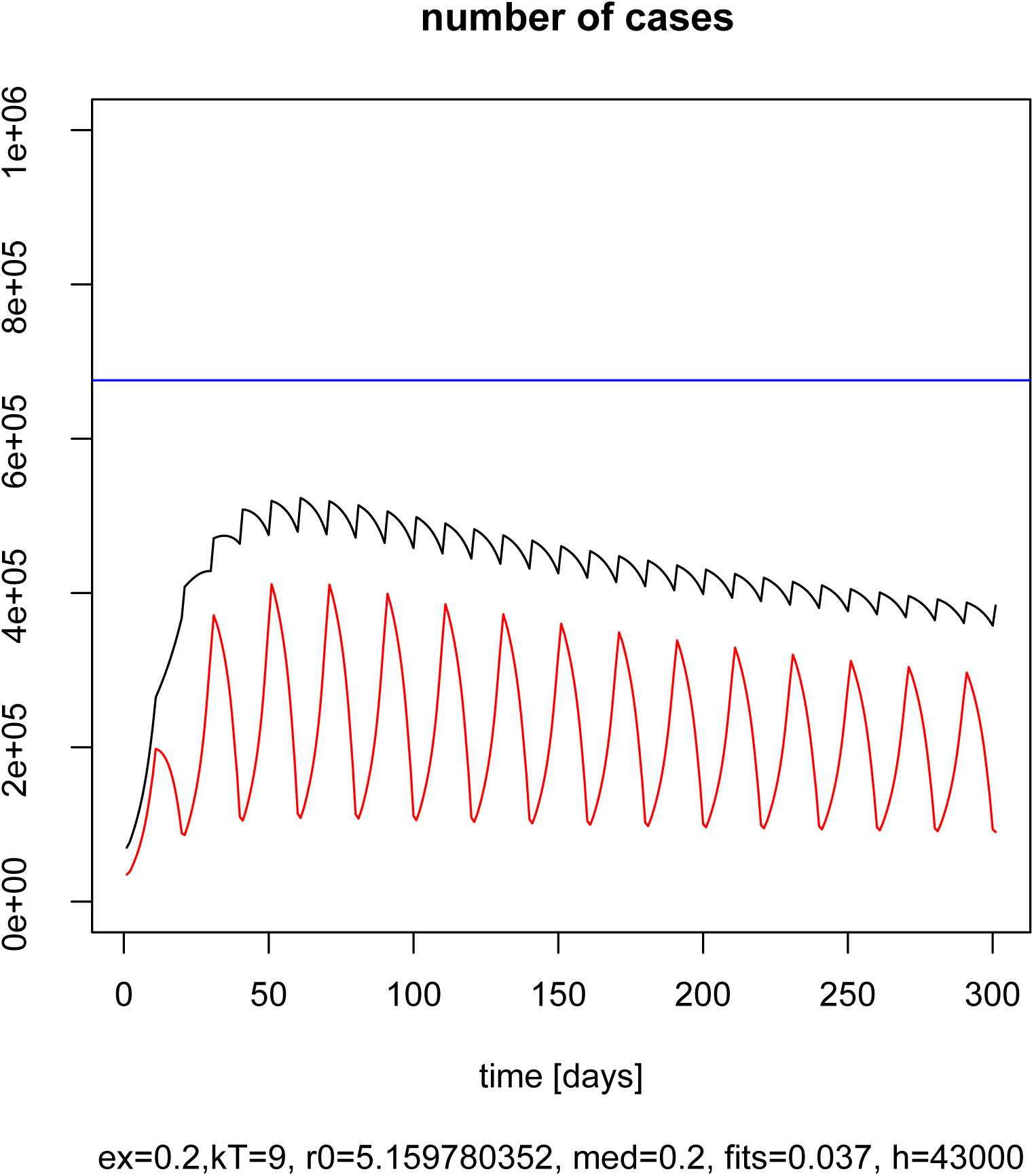

The two groups would alternate normal episodes and quarantine episodes in opposite directions. In the timeline on the left the red line shows one group and the black line portrays the sum of the two groups. Quarantine and normal episodes are mathematically defined in the same way as in the original model. An advantage of this option is the continuity of Germany economic output. Another advantage is better utilization of the country’s health care capacity thanks to smaller oscillations.

The problem with this solution is the high administrative burden in managing these two groups, in monitoring strategy compliance by the current quarantine group, assuring that branches of business are evenly distributed and simultaneously assigning all members of each household to the same group.

## Data Availability

All data are available on request.

### A1: Essential occupations in Germany

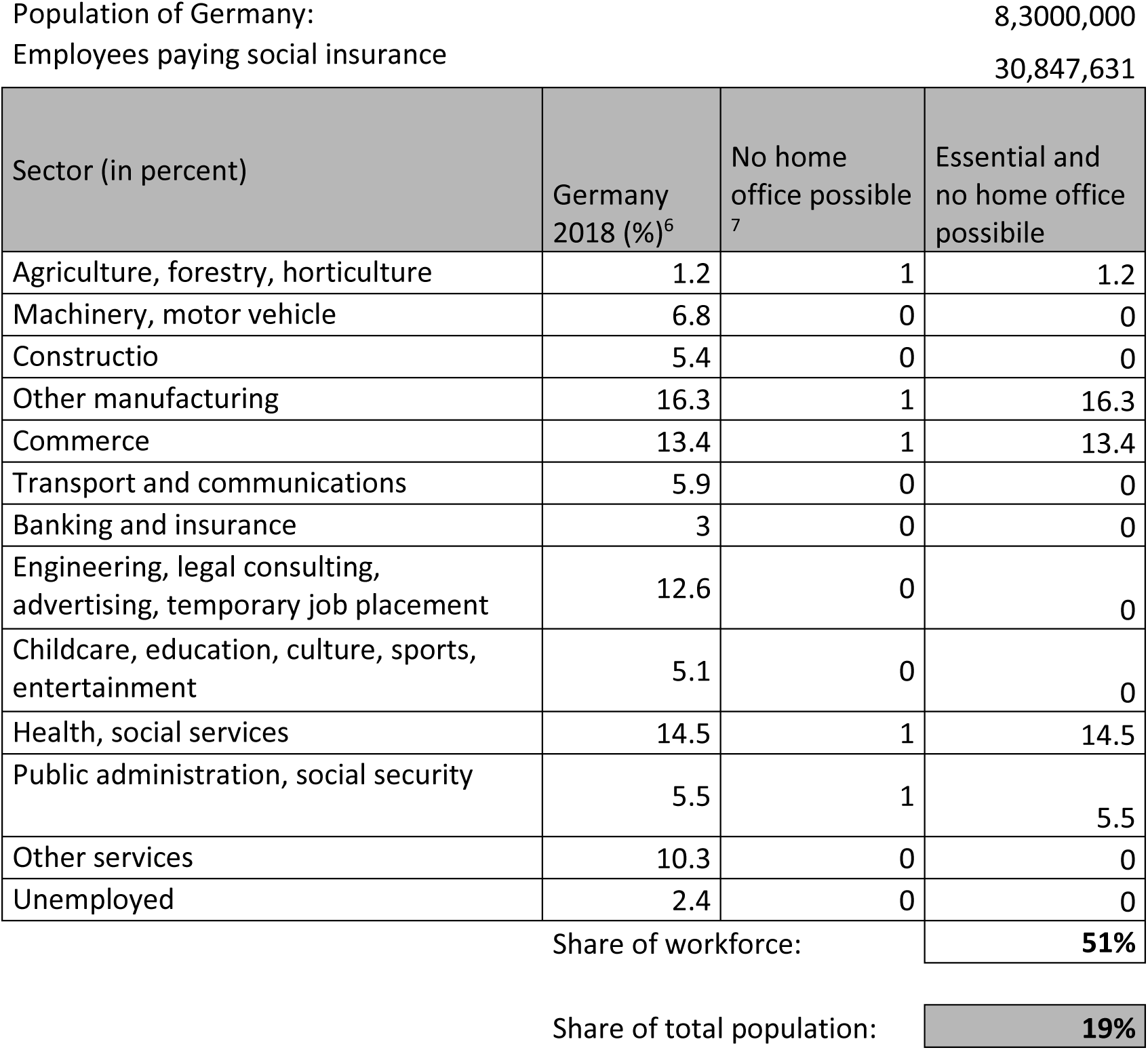

### A2: Estimation of COVID-19 cases requiring intensive care (IC) in Germany

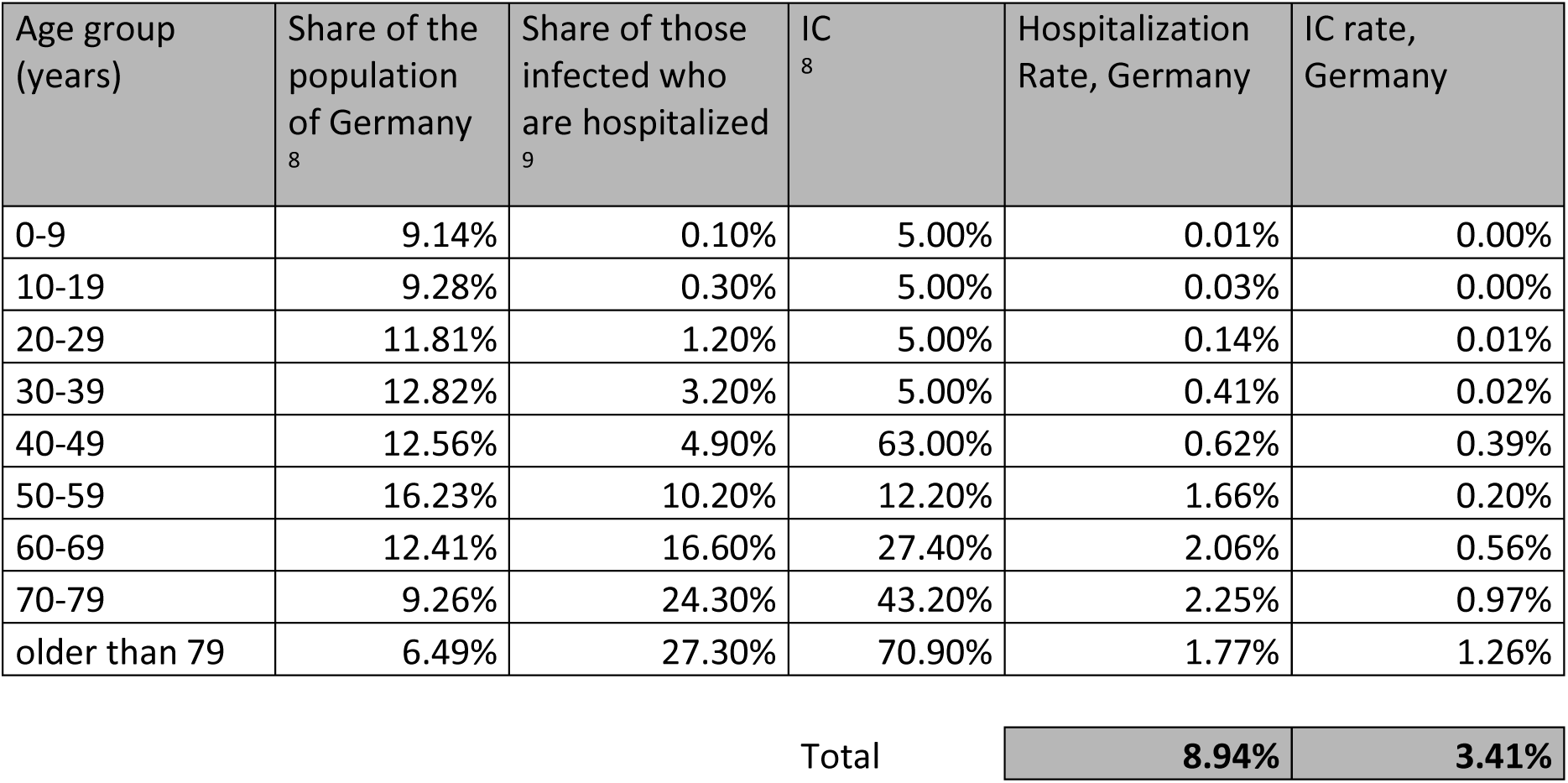

### A3: Calculation of the unfavorable parameter combination

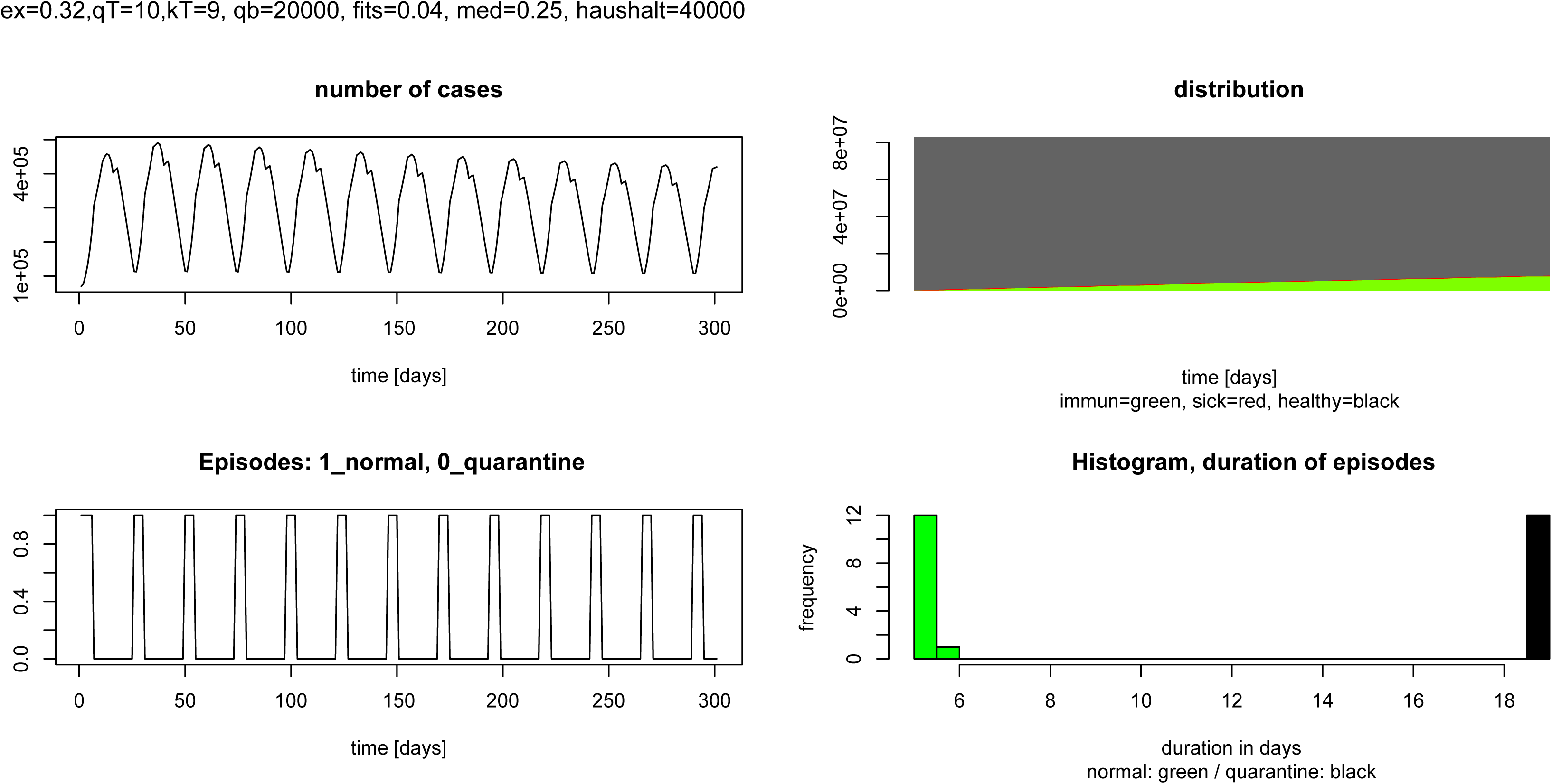

### A4: Calculation of the favorable parameter combination Nr.3

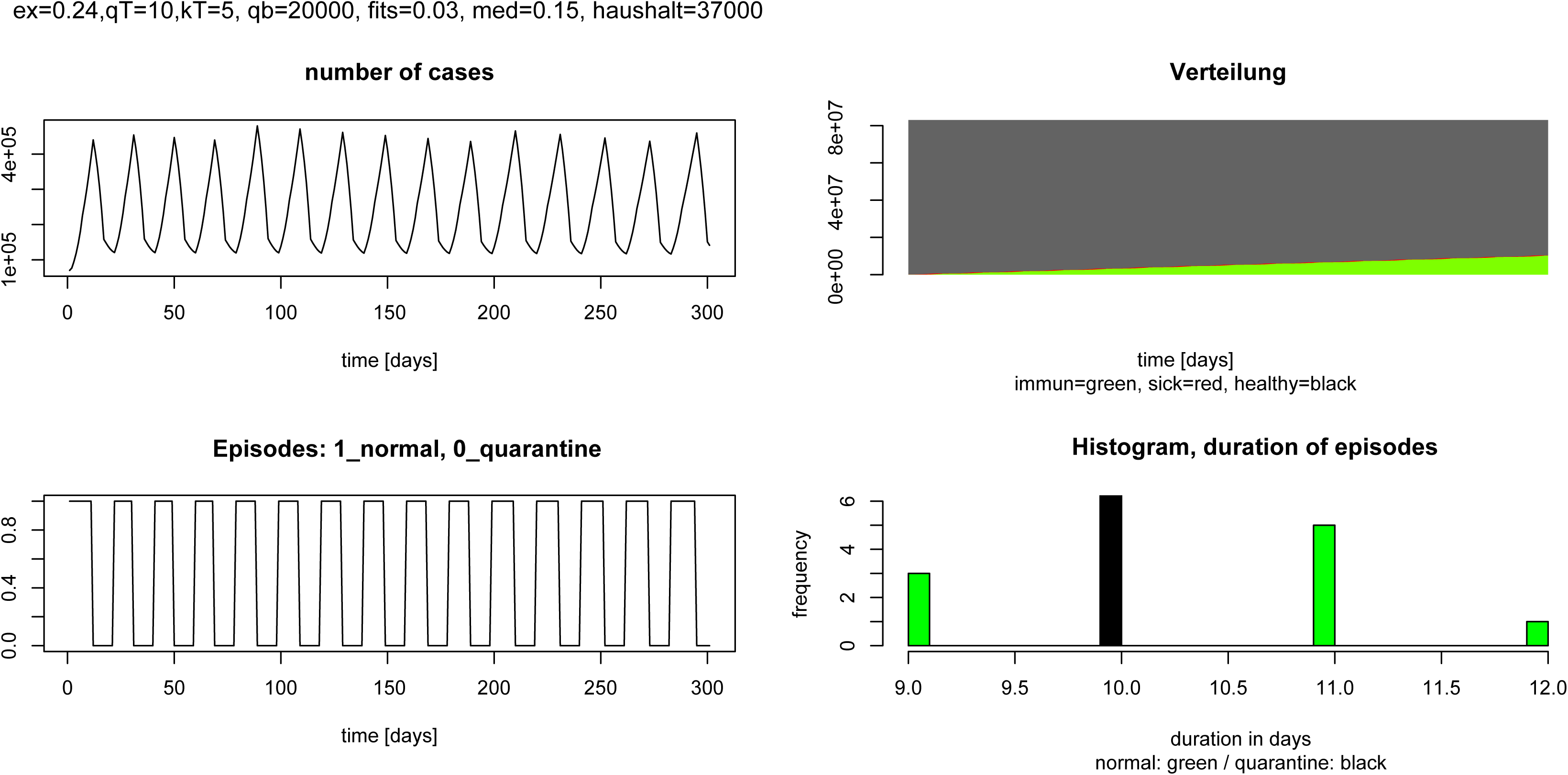

## Footnotes

1 https://www.rki.de/DE/Content/InfAZ/N/Neuartiges_Coronavirus/Steckbrief.html

2 Epidemiological Bulletin: https://www.rki.de/DE/Content/Infekt/EpidBull/Archiv/2020/Ausgaben/15_20.pdf?__blob=publicationFile

3 16 March 2020 Imperial College COVID-19 Response Team: DOI: https://doi.org/10.25561/7748

4 https://www.genesis.destatis.de/genesis/online?operation=abruftabelleBearbeiten&levelindex=1&levelid=1585082422492&auswahloperation=abruftabelleAuspraegungAuswaehlen&auswahlverzeichnis=ordnungsstruktur&auswahlziel=werteabruf&code=12411-0005&auswahltext=&werteabruf=starten#astructure

5 https://www.bpb.de/nachschlagen/zahlen-und-fakten/soziale-situation-in-deutschland/61587/haushalte-nach-zahl-der-personen

6 http://bisds.iab.de/Default.aspx?beruf=ABO®ion=1&qualifikation=0

7 Assumed

8 https://www-genesis.destatis.de/genesis/online?operation=abruftabelleBearbeiten&levelindex=1&levelid=1585082422492&auswahloperation=abruftabelleAuspraegungAuswaehlen&auswahlverzeichnis=ordnungsstruktur&auswahlziel=werteabruf&code=12411-0005&auswahltext=&werteabruf=starten#astructure

9 16 March 2020 Imperial College COVID-19 Response Team, DOI: https://doi.org/10.25561/77482

